# Tools for categorization of diagnostic codes in hospital data: Operationalizing CCSR into a patient data repository

**DOI:** 10.1101/2022.11.29.22282888

**Authors:** Sarah Malecki, Anne Loffler, Daniel Tamming, Michael Fralick, Shahmir Sohail, Jiamin Shi, Surain Roberts, Michael Colacci, Fahad Razak, Amol Verma

**Author notes:** Corresponding author **Correspondence To:** Amol Verma, Li Ka Shing Knowledge Institute St Michael’s Hospital 30 Bond Street, 14-077B Toronto, ON M5B 1W8. **Ethics Approval:** Research ethics board approval was obtained from the University Health Network (Toronto), Sunnybrook Health Sciences Centre (Toronto) and St. Michael’s Hospital (Toronto) through the integrated Clinical Trials Ontario platform, with St. Michael’s Hospital as the Board of Record (CTO project ID: 1394). Research ethics board approval was also obtained from Trillium Health Partners (Mississauga; REB# 742) and Mount Sinai Hospital (Toronto; REB# 15-0075-C).

## Abstract

**Background:** The Clinical Classification Software refined version (CCSR) is a tool to aggregate *International Classification of Diseases, 10th Revision, Clinical Modification/Procedure Coding System* (ICD-10-CM/PCS) diagnosis codes into clinically meaningful categories. ICD-10-CM/PCS codes are primarily used in the United States and the tool has not been optimized for use with other country-specific ICD-10 coding systems.

**Method:** We developed an automated procedure for mapping Canadian ICD-10 codes (ICD-10-CA) to CCSR categories using discharge diagnosis data from adult medical hospitalizations at 7 hospitals between Apr 1 2010 and Dec 31 2020, and manually validated the results.

**Results:** There were 383,972 Canadian hospital admissions with 5,186 distinct ICD-10 discharge diagnosis codes. Only 46.6% of ICD-10-CA codes could be mapped directly to CCSR categories. Our algorithm improved mapping of hospital codes to CCSR categories to 98.2%. Validation of the algorithm demonstrated a high degree of accuracy with strong interrater agreement (observed proportionate agreement of 0.98). The algorithm was critical for mapping the majority of diagnosis codes associated with heart failure (96.6%), neurocognitive disorders (96.0%), skin and subcutaneous tissue infections (97.2%), and epilepsy (92.5%).

**Conclusion:** Our algorithm for operationalizing CCSR into a patient data repository (https://github.com/GEMINI-Medicine/gemini-ccsr) has been validated for use with Canadian ICD-10 codes and may be useful to clinicians and researchers from diverse geographic locations.

## Introduction

The World Health Organization maintains the *International Classification of Diseases* (ICD) coding system to create a standard for recording data about human diseases.^1^ ICD codes support a wide range of healthcare applications, including administration of healthcare, quality measurement and reporting, payment, and health services research. ICD codes are developed based on a tree-like structure, with increasing characters responding to greater granularity of conditions. The 10^th^ revision (ICD-10) is now widely used, with more than 70,000 ICD-10 codes in some country-specific applications.^2^ The complexity of ICD-10 coding systems, however, poses an important challenge as clinical concepts (like disease conditions or symptomatic presentations) may be represented by numerous different ICD-10 codes, and individual ICD-10 codes may map to multiple clinical concepts.

The U.S. Agency for Healthcare Research and Quality sponsored the development of the Clinical Classifications Software Refined (CCSR), a tool that groups ICD-10 diagnosis codes into approximately 500 clinically meaningful categories.^2^ This type of tool facilitates a wide range of healthcare applications, including enabling easier reporting of healthcare utilization and outcomes related to various diseases^2-6^ and dimensionality reduction in statistical modeling ^7-11^. The CCSR was developed for use with ICD-10 *clinical modification/procedure coding system* (ICD-10-CM/PCS) codes, which are used in the United States. It is not known how well this tool works with different versions of ICD-10 coding systems that are used internationally.

The purpose of this study was to develop and validate an automated procedure for applying the CCSR tool in a Canadian hospital data repository.

## Methods

### Data Sources

For algorithm development and validation, we used data from the GEMINI retrospective cohort study.^12^ This initiative collects clinical and administrative data for adult medical hospitalizations from participating hospitals in Ontario, Canada. The administrative data include patient diagnoses which are mandatorily reported to the Canadian Institute for Health Information (CIHI).^13^ Diagnoses are recorded through manual chart review by trained chart abstractors, and up to 25 diagnoses are reported using the Canadian version of ICD-10 (ICD-10-CA).^14^

### Sample

We included all adults discharged from 7 hospitals between April 1, 2010 and October 31, 2019, where admission or discharge was from general internal medicine. Because the GEMINI study expanded its inclusion criteria beginning in 2019, we also included all adults discharged between November 1, 2019 and December 31, 2020, where admission was to any medical service (general internal medicine or subspecialties) or an intensive care unit. For each hospitalization, the most responsible diagnosis code was extracted.

### Developing a Procedure for Implementing CCSR with ICD-10-CA and algorithm validation

We developed an algorithm to automate the mapping of ICD-10-CA diagnosis codes to their corresponding CCSR category(s). The procedure relies on data in the official CCSR tool (v2020.3: https://www.hcup-us.ahrq.gov/toolssoftware/ccsr/ccsr_archive.jsp), which assigns each ICD-10-CM/PCS code to one or more (up to 5) CCSR categories. The existing mappings are then used to predict appropriate CCSR categories for ICD-10-CA codes based on their similarity to the ICD-10-CM/PCS codes. The algorithm was developed and implemented in Python. The source code is available at https://github.com/GEMINI-Medicine/gemini-ccsr. The automated mapping was followed by manual validation and adjustment by a subject matter expert (resident physician).

Each unique ICD-10-CA code was mapped according to the procedure illustrated in Figure 1. ICD-10-CA codes that existed in the official CCSR mapping file were mapped directly to their CCSR categories. If ICD-10-CA codes did not exist in the CCSR mapping file, an iterative procedure was used to match codes with closely related codes based on their hierarchical structure.

**Figure 1.**
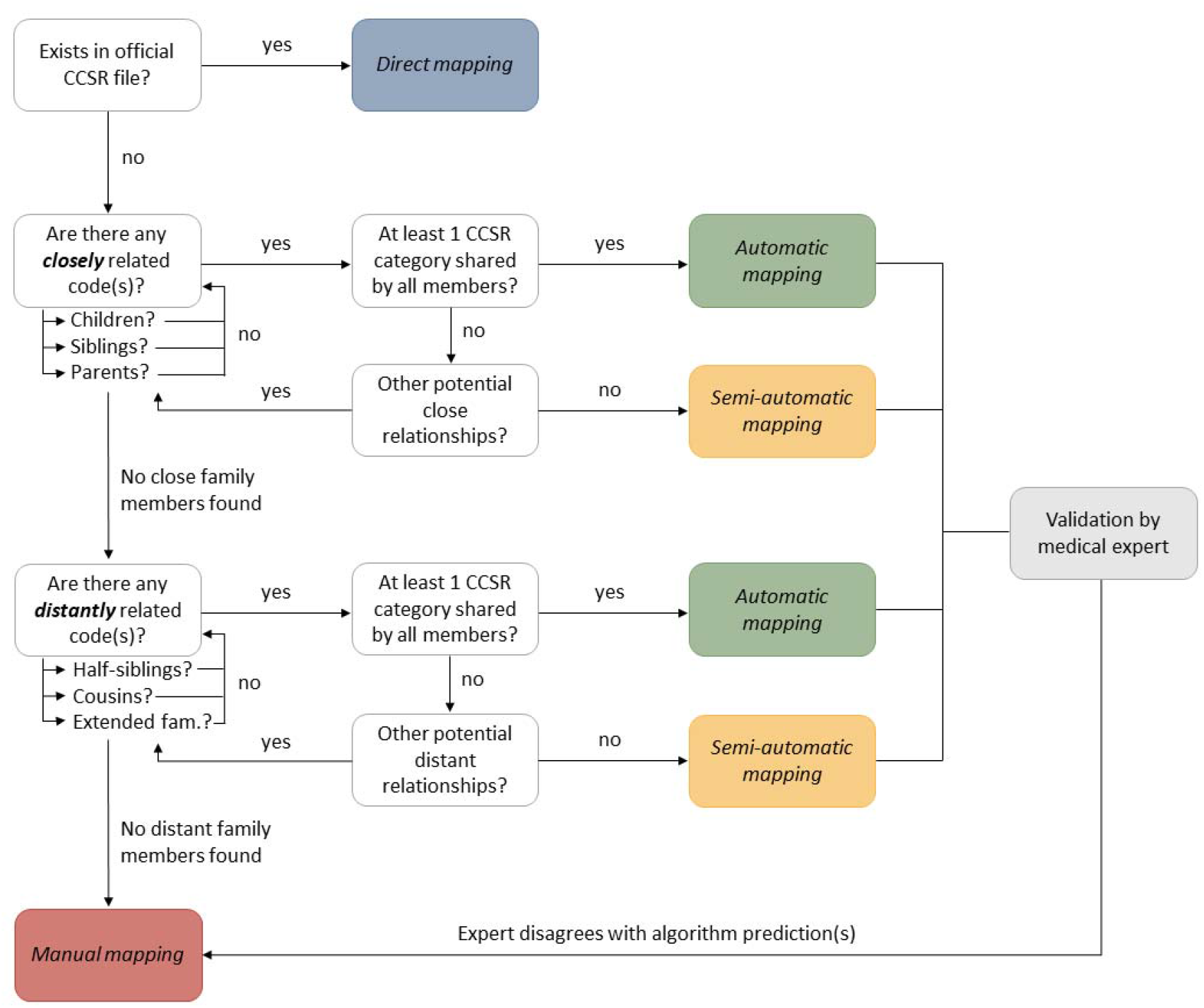
Schematic of mapping procedure. Each ICD-10-CA diagnosis code was mapped based on its similarity to existing diagnosis codes in the official CCSR file. If the queried code existed in the CCSR file, it could be mapped directly. Otherwise, the algorithm used an iterative procedure to search for similar codes that were either closely related (children/sibling/parents) or distantly related (half-siblings/cousins/extended family). If related codes were found, and they all shared at least one common CCSR category, the diagnosis code was mapped automatically. If the related codes did not all share a common CCSR category, all partially shared CCSR categories were returned as potential mappings (semi-automatic mapping). Finally, if no similar codes were found in the CCSR file, or if the medical expert(s) did not agree with any of the predicted CCSR categories, the diagnosis code was mapped manually.

“Children” codes were defined as ICD-10-CM codes that contain the queried ICD-10-CA code, but have one or more additional characters. For example, “A4181” is a child of “A418”. Conversely, “C880” is a “parent” of “C8808”. “Siblings” were defined as codes that have the same number of characters but differ in their last digit (e.g., “B485” and “B488”). The algorithm sequentially searched for children, sibling, and parent codes in the CCSR file. If related codes were found (e.g., siblings), the algorithm next checked whether *all* sibling codes shared at least one common CCSR category. If they did, the algorithm terminated and mapped the ICD-10-CA code to the categories that were shared by all of its sibling codes (*automatic mapping*). Automatically mapped ICD-10-CA codes were returned together with a description of the diagnosis code and its mapped CCSR category(s). Two subject matter experts (resident physicians) validated the mapping by independently confirming/rejecting the algorithm predictions, with the main goal of mapping codes to at least one suitable CCSR category. Alternative CCSR categories were only suggested if none of the predictions were accurate (*manual mapping*). In case of disagreement between the two reviewers, the mapping was resolved by a third expert.

If close code relationships were found, but none of them resulted in automatic mapping (i.e., no CCSR category was shared by all children/siblings/parents), the algorithm returned a list of all the CCSR categories that were partially shared by any closely related codes. One medical expert (SM) then chose any appropriate category(s) among the algorithm predictions (*semi-automatic mapping*), and occasionally suggested additional categories. In order to facilitate semi-automatic mapping, the algorithm output additionally provided a ranking based on the percentage of related codes that shared each candidate CCSR category. If none of the predicted categories were accurate, the expert manually mapped the diagnosis code to an appropriate CCSR category.

In case no closely related codes were identified, the algorithm checked for distantly related codes. First, the algorithm checked for half-siblings, which are nearby codes with the same number of characters that can differ on their last *two* digits as long as 1) the first three characters are identical and 2) the last two digits only differ by a distance of +/-9 (e.g., “E1165” is a half-sibling of “E1170”). Next, the algorithm checked for cousins, which are any codes that share the same first three characters, regardless of the remaining characters (e.g., “F010” and “F0151”). Finally, any diagnosis codes that only share the first two characters were classified as extended family (e.g., “A970” and “A91). Distant code relationships only resulted in automatic mapping if *all* half-siblings/cousins/extended family members shared a common CCSR category (strict criterion).

For codes that could not be mapped automatically, distant code relationships were also helpful in predicting multiple candidate CCSR categories that the expert could choose from (semi-automatic mapping), minimizing the need for fully manual mapping. Semi-automatic mappings that were based on distantly related codes were returned together with the percentage of diagnosis codes that shared each potential CCSR category. However, due to the potentially large number and variety of distantly related codes, we only included CCSR categories that were shared by the nearest existing distant relationship (i.e., half-siblings took priority over cousins, which took priority over extended family members).

Finally, if no closely/distantly related codes were found in the CCSR file, the diagnosis code was returned as unmapped and the medical expert chose a suitable category manually.

Once each diagnosis code was mapped and validated, a default CCSR category was chosen based on the following criteria: If a code was mapped to a single CCSR category, this category was automatically chosen as default CCSR. If a code was mapped to multiple CCSR categories, the algorithm checked whether a given combination of CCSR 1–5 categories existed in the official mapping file and then chose the corresponding default category accordingly. If the combination did not exist, the default CCSR category was chosen based on which one of the mapped categories was most frequently the default category among related codes.

### Statistical Analysis

To assess performance of our automated procedure, we report the percentage of ICD-10-CA codes that were mapped directly, automatically, semi-automatically, or manually. Mapping results are reported in two different ways: First, at the level of unique ICD-10-CA codes (regardless of how often they occurred in our cohort), which provides insights into the computational/manual effort required to map the individual diagnosis codes. Second, we report results at the level of hospitalizations (i.e., relative to the frequency of each diagnosis code in our cohort), which emphasizes the clinical relevance of our mapping procedure in a large cohort of hospital patients. We further report the accuracy of the algorithm in terms of approval of the predicted mappings by the medical experts. Finally, to assess the impact of this procedure on clinical findings, we report mapping performance separately for each of the 30 most common CCSR categories in our cohort.

## Results

Baseline characteristics for the GEMINI cohort are displayed in Table 1. The cohort comprised 383,972 hospital admissions. 51.2% of admissions were for men, and the median age was 71 (IQR 56-83). In this cohort there were 5,186 distinct ICD-10 discharge diagnosis codes.

**Table 1.**
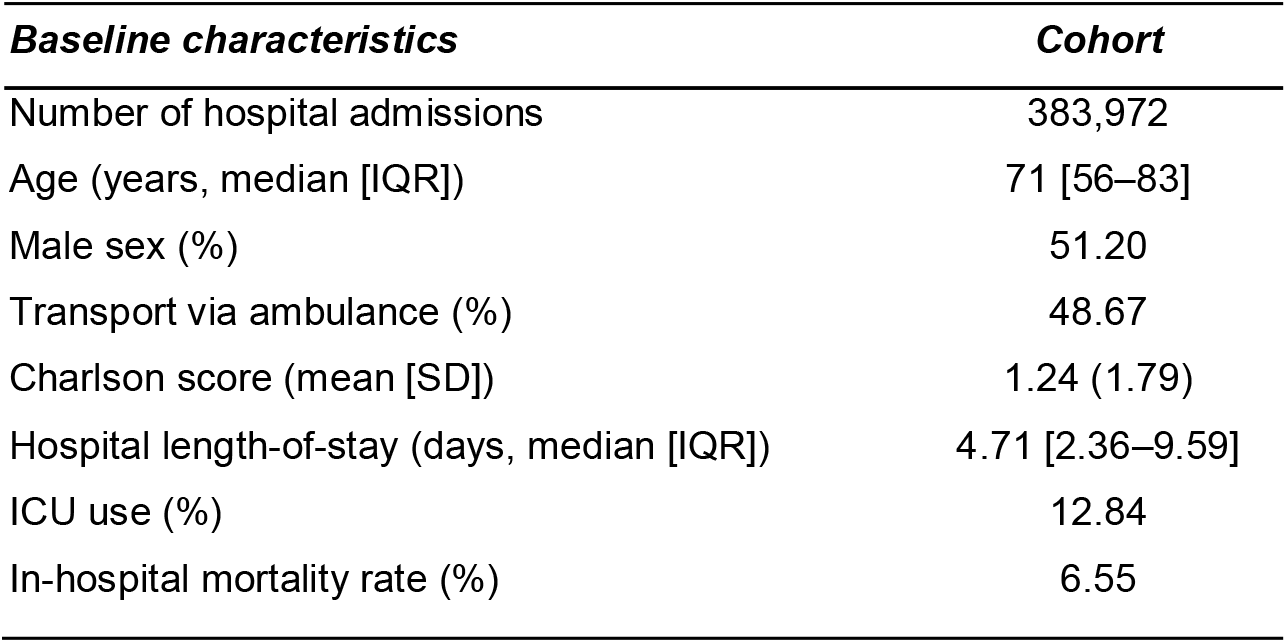
Characteristics of GEMINI hospitalizations

| **Baseline characteristics** | **Cohort** |
| --- | --- |
| Number of hospital admissions | 383,972 |
| Age (years, median [IQR]) | 71 [56–83] |
| Male sex (%) | 51.20 |
| Transport via ambulance (%) | 48.67 |
| Charlson score (mean [SD]) | 1.24 (1.79) |
| Hospital length-of-stay (days, median [IQR]) | 4.71 [2.36–9.59] |
| ICU use (%) | 12.84 |
| In-hospital mortality rate (%) | 6.55 |

### Performance and Validation of Automated Procedure in GEMINI data

Using the CCSR tool alone on the GEMINI dataset, we were able to directly map 46.6% of unique ICD-10-CA diagnostic codes to CCSR categories (Fig. 2A, left), representing 56.5% of all hospitalizations (Fig. 2A, right). An additional 31.3% of unique diagnosis codes (33.4% of hospitalizations) were successfully mapped automatically, and 20.3% (9.14% of hospitalizations) were mapped semi-automatically. The remaining 1.79% of diagnosis codes (0.88% of hospitalizations) were mapped manually. The manually mapped codes largely reflected diagnosis codes in which the medical experts disagreed with the automatically/semi-automatically derived mappings, but they also included a single ICD-10-CA code (“E90”; Nutritional and metabolic disorders in diseases classified elsewhere) for which the algorithm did not provide any predictions due to no existing related codes in the CCSR file.

**Figure 2.**
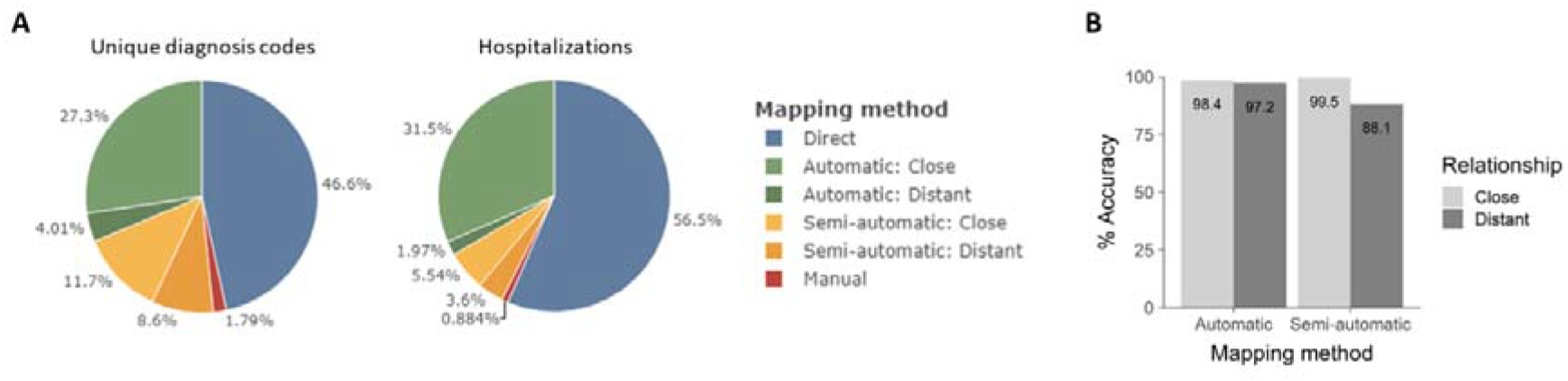
Performance of mapping algorithm. A) Percentage of different mapping methods at the level of unique diagnosis codes (left) and hospitalizations (right). For automatic (green) and semi-automatic (yellow) mapping, shading indicates whether mappings were derived based on close (light) or distant (dark) code relationships. B) Accuracy of automatic and semi-automatic mapping, separately for diagnosis codes that were mapped based on close (light grey) vs. distant (dark grey) code relationships. Accuracy reflects the percentage of ICD-10-CA codes for which the expert(s) agreed with at least one of the predicted CCSR categories.

Among automatic mappings, the majority of unique diagnosis codes (87.2%) were mapped based on close code relationships, with children codes being the most frequent category accounting for 54.1% of automatically mapped codes. Distantly-related codes only made up a small portion (12.8%) of automatic mappings because half-siblings/cousins/extended family codes were less likely to all share a common CCSR category. However, distant code relationships substantially improved our ability to predict a list of potential mappings based on partially shared CCSR categories (semi-automatic mapping). Of all codes that were mapped semi-automatically, 42.4% were mapped based on distant code relationships, with cousins being the most frequent type of distant codes (86.2%). Overall, distant code relationships enabled us to map 12.6% of unique ICD-10-CA codes, which otherwise would have had to be mapped completely manually.

At the level of hospitalizations, we observed that distant code relationships only accounted for 5.88% of mapped diagnoses. More generally, we observed that higher mapping difficulty was associated with less frequent diagnoses. That is, mappings that were based on distant code relationships and/or involved more manual work (semi-automatic/manual mapping) accounted for smaller portions of the data when we analyzed diagnosis codes at the level of hospitalizations (Fig. 2A, right) compared to the level of unique diagnosis codes (Fig. 2A, left). By contrast, the proportion of codes that could be mapped directly/automatically was larger at the level of hospitalizations, meaning more frequent diagnosis codes were associated with easier, more automated mapping mechanisms (also see Fig. 3 below).

**Figure 3.**
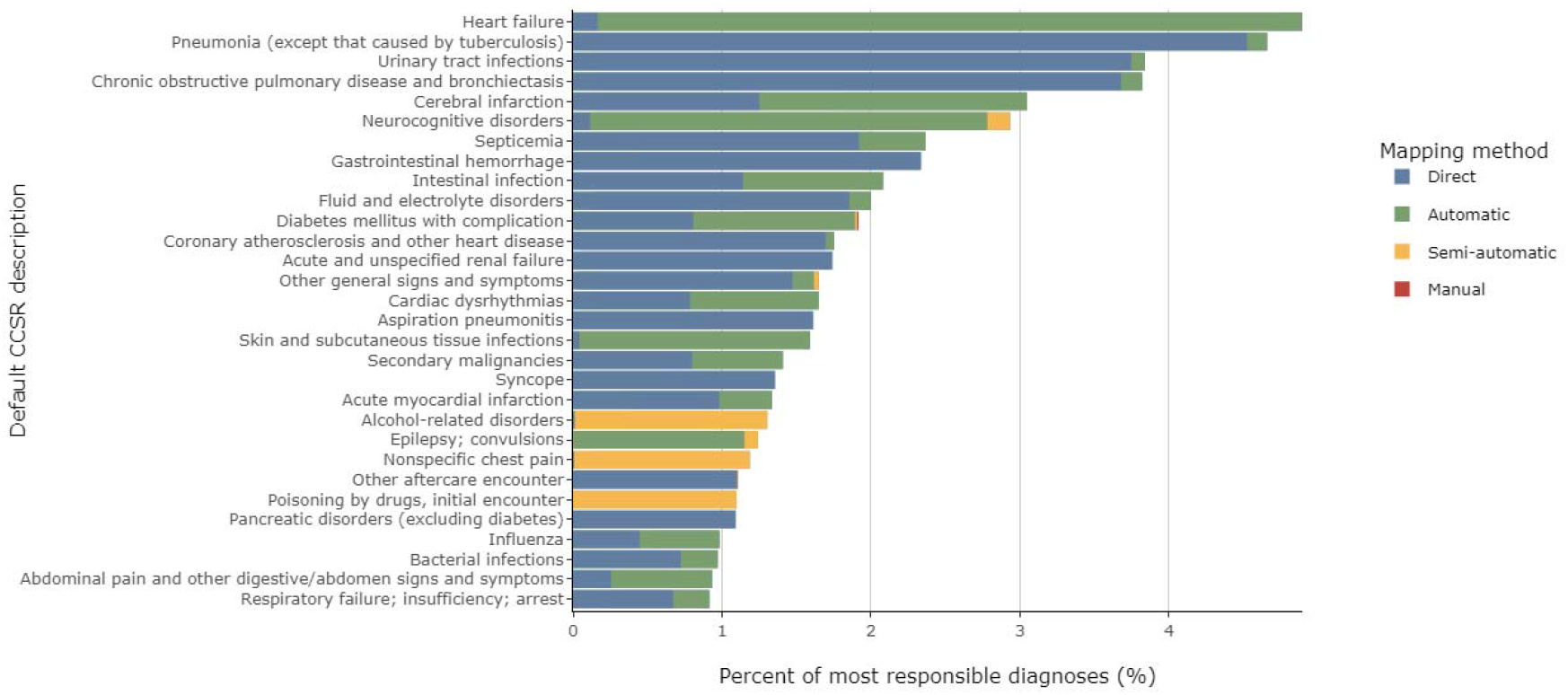
Mapping methods for the 30 most common default CCSR categories. For each CCSR category, colors indicate the proportion of diagnosis codes that were mapped directly (blue), automatically (green), semi-automatically (yellow), or manually (red). For illustration purposes, CCSR categories were sorted according to their prevalence in the GEMINI cohort, with the most frequent category at the top.

Finally, we analyzed the accuracy of the mapping algorithm. Although the medical experts had to correct some of the predicted mappings, approval rate was very high overall. For automatically mapped codes, both reviewers agreed with 1,750/1,804 (97.01%) of unique diagnosis codes (Supplemental Table 1). Both experts rejected 24 (1.33%) automatic algorithm predictions. Overall, analysis of inter-rater reliability showed a high level of agreement, with an observed proportionate agreement of 0.98. Results shown in Figure 2B illustrate the accuracy of automatic mappings, separately for close vs. distant code relationships, after disagreements were resolved by a third expert. For semi-automatic mappings, the single reviewer approved at least one of the predicted CCSR categories for 1,053/1,145 (92.0%) of unique diagnosis codes. Approval rate was lowest for semiautomatic mappings that were derived based on distantly related codes (Fig. 2B).

### Algorithm application to Canadian hospital data

Finally, we assessed mapping performance for individual CCSR categories. Figure 3 shows the breakdown of direct versus algorithm-based mapping for the 30 most common default CCSR categories in our cohort. Applying the matching algorithm substantially changes the proportion of patients identified with many common conditions (Figure 3).

## Discussion

We designed a practical algorithm for applying the CCSR tool to Canadian hospital data, which was validated by medical experts and resulted in improved mapping of diagnostic codes from 46.6% to 98.2%. The algorithm was especially important for capturing common admission diagnostic categories in general internal medicine such as heart failure and dementia. Clinicians and researchers from different geographic locations and institutions may find our method useful for applying CCSR categorization to their own hospital data.

Application of our algorithm in Canadian data allowed us to achieve and surpass the rate of mapping in a US report using ICD-10 CM codes, where 92% of hospital codes extracted from a mortality database were directly mapped to a CCSR category.^3^ Our results illustrate how direct application of a US-based tool in another country can be improved to achieve excellent performance.

Our algorithm combined simple rules with an iterative process to achieve a high percentage of mapped codes. Drawing on close code relationships (parents and children) allowed us to map the majority of codes, similar to a recent study in Singapore showing a high degree of success in conversion of ICD-10 codes in the country of origin to US ICD-9 codes with a simple algorithm of creating child (addition of 0s) or parent (truncation) codes. The converted codes were then able to be directly mapped to CCS categories.^15^ Our results also show that expanding the algorithm to consider more distant code relationships (e.g. cousins, half siblings) can be helpful for semi-automatic processing of a smaller proportion of less frequent codes that would otherwise need to be mapped completely manually.

The accuracy of our CCSR categorization algorithm is supported by a high level of agreement among expert reviewers (98% agreement) and strengthened by consistency in the most common diagnostic categories compared to the earlier, more simplistic categorization tool CCS in our cohort.^16^ The top four most common diagnoses using both methods were heart failure, pneumonia, COPD and UTI.

The clinical utility of our algorithm is demonstrated by the fact that some common diagnoses in Canadian hospital data would not have been possible to map otherwise. Our algorithm was critical for mapping the majority of diagnosis codes associated with heart failure (96.6%), neurocognitive disorders (96.0%), skin and subcutaneous tissue infections (97.2%), and epilepsy (92.5%). Clinical utility of this algorithm is expected in other countries as well, given that other countries have implemented their own specific version of ICD-10.^17^

The major limitations of our work include that this study was developed and validated only in adult medical inpatients at 7 hospitals. Extension to surgical and other non-medical inpatients or outpatient populations requires further validation, although there is no reason to suspect that the automated and semi-automated matching procedure would work any less well in these other settings. It is also worth noting that the validation of CCSR category assignment was not blinded, i.e. the subject matter experts were reacting to suggested matches. It is possible that they would have selected different category matches if presented the ICD-10-CA codes without any suggestions. The suggested matching procedure was designed to minimize manual work and facilitate expert review, and our work demonstrates that it was able to successfully do so.

Overall, our algorithm operationalizing CCSR into a patient data repository is simple, freely available, and offers a way to substantially improve direct mapping using the available US-based online CCSR tool. This offers a way to explore disease categories in Canadian hospitalization data and can be easily applied and validated in other settings around the world.

## Data Availability

Data from this manuscript can be accessed upon request to the corresponding author, to the extent that is possible in compliance with local research ethics board requirements and data sharing agreements.

## Supplementary Material

### Supplementary Tables

**Table S1.**
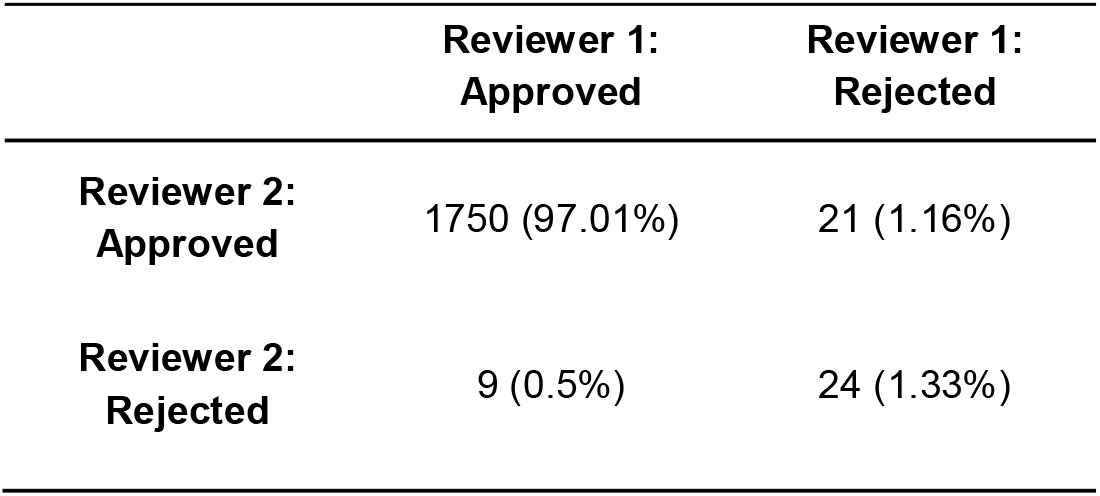
Reviewer approval/rejection rate for automatically mapped codes.

